# Efficacy of lightweight Vision Transformers in diagnosis of pneumonia

**DOI:** 10.1101/2024.10.24.24316057

**Authors:** Muhammad Tayyeb Bukhari

## Abstract

Pneumonia is one of the leading causes of death in children under five, particularly in resource-limited settings. The timely and accurate detection of pneumonia, often conducted through chest X-rays, remains a challenge due to the scarcity of trained professionals and the limitations of traditional diagnostic methods. In recent years, Artificial Intelligence (AI) models, especially Convolutional Neural Networks (CNNs), have been increasingly applied to automate pneumonia detection. However, CNN models are often computationally expensive and lack the ability to capture long-range dependencies in images, limiting their efficacy in certain medical applications. To address these limitations, lightweight hybrid models such as Vision Transformers (ViTs), which combine the strengths of CNNs and transformers, offer a promising solution. This study compares the efficacy of two lightweight CNNs (EfficientNet Lite0 and MobileNetV3 Large) with two hybrid ViTs (MobileViT Small and EfficientFormerV2 S0) for pneumonia detection. The models were evaluated on a publicly available chest X-ray dataset using metrics such as accuracy, F1 score, precision, and recall. Results show that the hybrid models, particularly MobileViT Small, outperformed their CNN counterparts in both accuracy (97.50%) and F1 score (0.9664), demonstrating the potential of ViT-based models for medical imaging tasks. The findings suggest that hybrid models provide superior recall, reducing false negatives, which is crucial for medical diagnostics. Further research should focus on optimizing these hybrid models to improve computational efficiency while maintaining high diagnostic performance.

## 2 Introduction

In 2019, pneumonia killed 740000+ children under the age of five respectively making it one of the leading causes of death in vulnerable populations. Pneumonia is an acute respiratory infection due to a viral or bacterial pathogen which targets the lungs, the alveoli in particular filling them with pus and fluid. Typical symptoms include cough, shortness of breath, chest pain, fatigue, and fever (World Health Organization [WHO], 2022).

In regions with limited healthcare resources, the timely and accurate diagnosis of pneumonia poses significant challenges due to a shortage of trained professionals and inadequate facilities (Simkovich et al., 2021). Pneumonia detection is commonly done using chest x-rays however traditional methods are time-consuming and prone to human error (Alapat et al., 2022). This is where automated image analysis comes into play, Artificial Intelligence (AI) tools, such as Convolutional Neural Networks (CNNs), are increasingly being applied to improve accuracy in detecting pneumonia, particularly in resource-limited settings (An et al., 2024).

Many powerful AI models exist but since they are computationally expensive, so they are not suited for resource constrained environments (Jia et al., 2023). In cases like this, lightweight models like MobileNet are employed which balance efficiency and accuracy (Trivedi & Gupta, 2021). The purpose of this study is to compare the efficacy of lightweight hybrid Vision Transformers (ViTs) and CNNs for pneumonia detection, focusing on both performance and efficiency, with the hypothesis that hybrid ViTs consisting of elements of CNNs and ViTs will outperform traditional CNNs.

## 3 Materials and Methods

### 3.1 Dataset

The dataset used was the Mendeley chest X-ray pneumonia dataset (Kermany et al., 2018) and can be accessed <u>here</u>. The train images, which totalled to 5216 images (of which 3875 were labelled as pneumonia and 1341 were labelled as normal), were used for training, validating, and testing. The images were split into training, validation, and test datasets in the ratio 7:2:1.

### 3.2 Data Augmentation

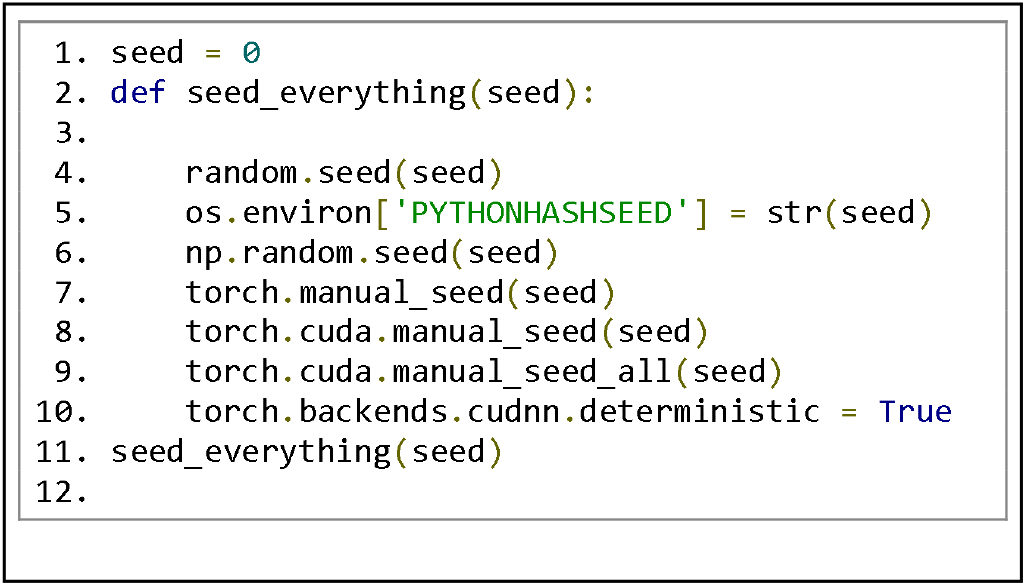

The seed was set to 0 to ensure reproducibility using the following code:

#### 3.2.1 Training dataset augmentation

Various augmentations from the albumentations (Buslaev et al., 2020) library were implemented to ensure model generalisation by introducing variability in the images. The augmentations applied are as follows:

- Rotation: The images were rotated to up to ±180° with a probability of 0.5.
- Affine: Scaling between 0.9 to 1.1, translation up to 10%, and shear between -2 and 2 were applied with a probability of 0.5.
- Flipping: Images were randomly flipped horizontally and vertically with a 50% probability for both.
- Resizing: All images were resized to 224×244 pixels
- Normalization: The RGB values of pixels were mapped to be in the range 0 to 1.

#### 3.2.2 Validation and Test dataset augmentation

The transformation pipeline for the validation dataset and test dataset was simpler and was as follows:

- Resizing: Images were resized to 224×224 pixels.
- Normalization: RGB values were normalized to be in the range 0 to 1.

### 3.3 MODEL ARCHITECTURES

Four lightweight image classification models were employed in this study:

- EfficientNet Lite0
- MobileNetV3 Large
- MobileViT Small
- EfficientFormerV2 S0

#### 3.3.1 EfficientNet Lite0

A scaled-down version of the original EfficientNet model, EfficientNet Lite0 was created with mobile and edge devices in mind. It makes use of MBConv layers, which combine squeeze-and-excitation blocks with depthwise separable convolutions. By keeping performance high while lowering the number of parameters, these layers strike a balance between accuracy and efficiency. The model employs a compound scaling method that uniformly scales depth, width, and resolution of the network based on a predefined factor. This enables the model to perform well across various tasks with fewer computations (Tan, & Le, 2019).

#### 3.3.2 MobileNetV3 Large

MobileNetV3 Large is intended for use by mobile devices with constrained computational resources. It uses squeeze-and-excitation layers, inverted residuals, and depthwise separable convolutions. For increased accuracy at the lowest possible computational cost, it also makes use of hard-swish and swish activation functions. MobileNetV3 further optimizes performance by using Neural Architecture Search (NAS) to identify the best configurations of layers. Higher accuracy is given priority in the large version, but the lightweight design is kept (Howard et al., 2019).

#### 3.3.3 MobileViT Small

The goal of MobileViT, a ViT-CNN hybrid model, is to integrate the local feature extraction powers of CNNs with the globally receptive field of Vision Transformers (ViT). MobileViT combines CNNs that extract local features with transformer encoders to capture long-range dependencies in the image. Because of this combination, MobileViT offers a balance between accuracy and efficiency that is optimal for mobile applications (Mehta & Rastegari, 2021).

#### 3.3.4 EfficientFormerV2 S0

An effective vision transformer for quick inference on edge devices is EfficientFormerV2. To achieve high performance on image classification tasks, it combines the simplicity of depthwise separable convolutions with the efficiency of transformers. Memory and computational expenses are decreased by EfficientFormerV2 models, such as the S0 variant, by using an optimized transformer block and a hierarchical design. MLP (Multi-Layer Perceptron) blocks and multi-head self-attention mechanisms have been integrated into the architecture of EfficientFormerV2. One of the smallest models, the S0 version was created especially for mobile and edge applications (Li et al., 2022).

### 3.4 Training and Optimization

All models were trained for 30 epochs using the following setup:

- Optimizer: The Adam optimizer with a learning rate of 1e-4 and a weight decay of 1e-5 was used to minimize the loss.
- Loss function: The loss function used as the criterion was Binary Cross-Entropy with Logits Loss as it was suited for this binary classification problem.
- Scheduler: ReduceLROnPlateau was employed which would reduce the learning rate by a factor of 0.1 when the validation loss would plateau.

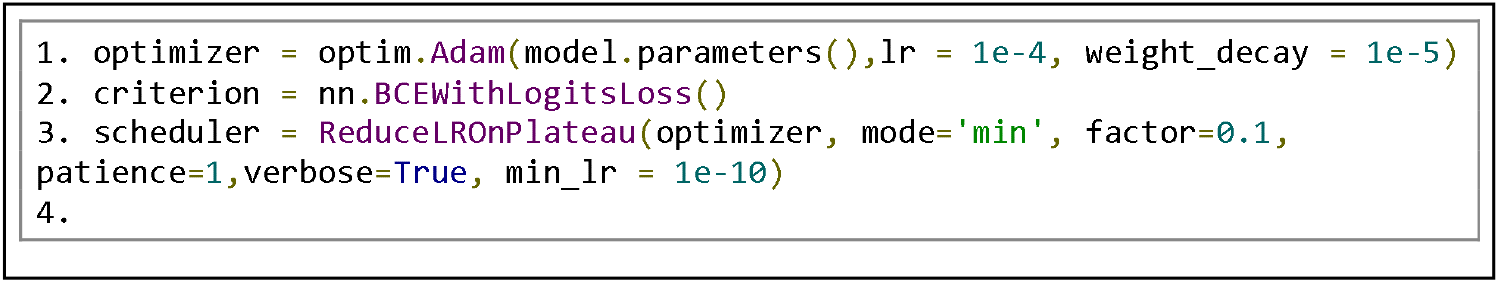

The following code was used to configure the optimizer, the loss function, and the scheduler:

In each epoch, the model was trained on the training dataset and then evaluated on the validation dataset. The validation loss was used to update the scheduler to ensure that the learning rate was reduced when validation loss would stop decreasing.

### 3.5 Evaluation Metrics

After training each model was evaluated on the test set using the following metrics:

- Accuracy score: the proportion of correctly predicted labels out of the total.
- F1 score: the harmonic mean of precision and recall.
- Precision score: the ratio of true positives to the sum of true positives and false positives.
- Recall score: the ratio of true positives to the sum of true positives and false positives.

The metrics were calculated using the methods provided in the scikit-learn library which required a list of the predicted labels and a list of the true labels (Pedregosa et al., 2011).

## 4 Results

The following results were obtained from the evaluation of the models on the test dataset and computation of the above-mentioned metrics for each of the four models.

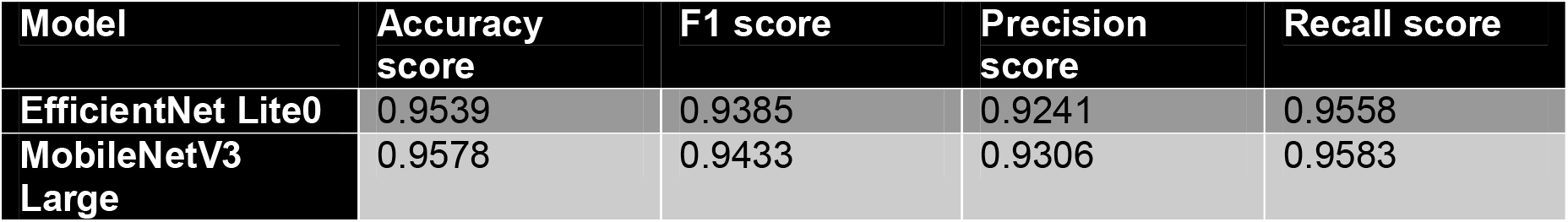
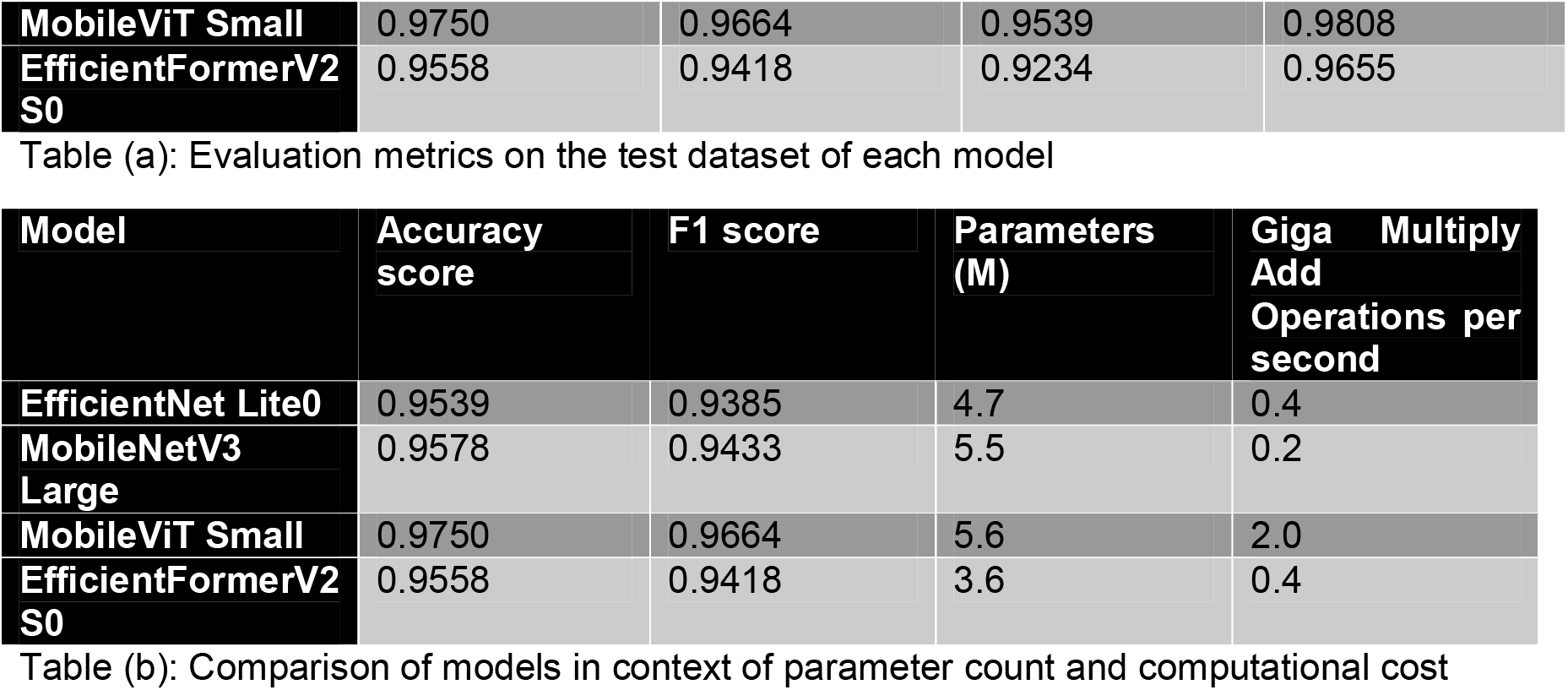

## 5 Discussion

### 5.1 Comparison of CNN and Hybrid ViT models

The results reveal that while both the CNN models and the hybrid ViT models performed well on the test dataset, both the hybrid ViTs performed better than their CNN counterparts in terms of accuracy score and F1 score. The best performing models was MobileViT Small with an accuracy score of 0.9750 and an F1 score of 0.9664, performing better than the other models in all metrics used in the study. The best performing CNN model was MobileNetV3 Large, achieving an accuracy score of 0.9578 and an F1 score of 0.9433 but fell short in recall score in comparison with the hybrid ViT models.

The CNN models rely heavily on local feature extraction through convolutional layers, which makes them efficient in identifying localized patterns within an image, such as the texture of lung tissues (Tan, & Le, 2019) (Howard et al., 2019). However, their ability to capture long-range dependencies across the image is limited, which could explain why they underperformed compared to hybrid models like MobileViT Small, which integrate both local and global feature extraction. This blend enables hybrid models to capture more complex relationships across the entire X-ray image, which is crucial in medical diagnostics where subtle, distributed patterns may indicate disease.

EfficientFormerV2 S0 was outperformed by both MobileNetV3 Large and MobileViT Small in terms of accuracy score and F1 score, achieving an accuracy score and F1 score of 0.9558 and 0.9418, respectively. It did however have a low GMACs value of 0.4 (Li et al., 2022) while attaining a high recall score of 0.9655, outperforming both CNN models in this regard. This suggests that transformer components in hybrid architectures significantly enhance performance by allowing the model to consider the entire image context and produce fewer false negatives.

### 5.2 Practical implications and further research

The results highlight that lightweight hybrid ViT models, such as MobileViT Small and EfficientFormerV2 S0, exhibit superior recall compared to lightweight CNN models, which is critical in medical applications like pneumonia detection, where false negatives can lead to missed diagnoses and delayed treatment. This makes hybrid models particularly valuable for preventing underdiagnosis, ensuring that more cases of pneumonia are correctly identified, even in resource-constrained settings. However, CNN models like MobileNetV3 Large still performed strongly, particularly in terms of computational efficiency, indicating that CNNs are not far behind and continue to be viable options for real-time applications where model complexity needs to be minimized.

Further research should focus on optimizing hybrid models to reduce their computational demands while maintaining high recall. Additionally, exploring hybrid architectures in diverse medical imaging tasks, along with investigating how CNNs can be enhanced to capture long-range dependencies, would provide valuable insights into creating more efficient and accurate diagnostic tools.

## Data Availability

All data produced in the present work are contained in the manuscript

https://doi.org/10.17632/rscbjbr9sj.2

## REFERENCES

World Health Organization. (2022). Pneumonia. https://www.who.int/news-room/fact-sheets/detail/pneumonia

Simkovich, S. M., Underhill, L. J., Kirby, M. A., Crocker, M. E., Goodman, D., McCracken, J. P., Thompson, L. M., Diaz-Artiga, A., Castañaza-Gonzalez, A., Garg, S. S., Balakrishnan, K., Thangavel, G., Rosa, G., Peel, J. L., Clasen, T. F., McCollum, E. D., Checkley, W., & HAPIN Investigators (2022). Resources and Geographic Access to Care for Severe Pediatric Pneumonia in Four Resource-limited Settings. American journal of respiratory and critical care medicine, 205(2), 183–197. 10.1164/rccm.202104-1013OC

Alapat, D. J., Menon, M. V., & Ashok, S. (2022). A Review on Detection of Pneumonia in Chest X-ray Images Using Neural Networks. Journal of biomedical physics & engineering, 12(6), 551–558. 10.31661/jbpe.v0i0.2202-1461

An, Q., Chen, W., & Shao, W. (2024). A Deep Convolutional Neural Network for Pneumonia Detection in X-ray Images with Attention Ensemble. Diagnostics (Basel, Switzerland), 14(4), 390. 10.3390/diagnostics14040390

Jia, Z., Chen, J., Xu, X., Kheir, J., Hu, J., Xiao, H., Peng, S., Hu, X. S., Chen, D., & Shi, Y. (2023). The importance of resource awareness in artificial intelligence for healthcare. Nature Machine Intelligence, 5(7), 687–698. 10.1038/s42256-023-00670-0

Trivedi, M., & Gupta, A. (2021). A lightweight deep learning architecture for the automatic detection of pneumonia using chest X-ray images. Multimedia Tools and Applications, 81(4), 5515–5536. 10.1007/s11042-021-11807-x

Kermany, D., Zhang, K., & Goldbaum, M. (2018). Labeled Optical Coherence Tomography (OCT) and chest X-Ray images for classification [Dataset]. In Mendeley Data. 10.17632/rscbjbr9sj.2

Buslaev, A., Iglovikov, V. I., Khvedchenya, E., Parinov, A., Druzhinin, M., & Kalinin, A. A. (2020). Albumentations: Fast and Flexible Image Augmentations. Information, 11(2), 125. 10.3390/info11020125

Tan, M., & Le, Q. V. (2019, May 28). EfficientNet: Rethinking Model Scaling for Convolutional Neural Networks. arXiv.org. 10.48550/arXiv.1905.11946

Howard, A., Sandler, M., Chu, G., Chen, L., Chen, B., Tan, M., Wang, W., Zhu, Y., Pang, R., Vasudevan, V., Le, Q. V., & Adam, H. (2019). Searching for MobileNetV3. arXiv (Cornell University). 10.48550/arxiv.1905.02244

Mehta, S., & Rastegari, M. (2021). MobileViT: Light-weight, General-purpose, and Mobile-friendly Vision Transformer. arXiv (Cornell University). 10.48550/arxiv.2110.02178

Li, Y., Hu, J., Wen, Y., Evangelidis, G., Salahi, K., Wang, Y., Tulyakov, S., & Ren, J. (2022). Rethinking Vision Transformers for MobileNet Size and Speed. arXiv (Cornell University). 10.48550/arxiv.2212.08059

Pedregosa, F., Varoquaux, G., Gramfort, A., Michel, V., Thirion, B., Grisel, O., Blondel, M., Prettenhofer, P., Weiss, R., Dubourg, V., Vanderplas, J., Passos, A., Cournapeau, D., Brucher, M., Perrot, M., & Duchesnay, É. (2011). Scikit-learn: Machine Learning in Python. Journal of Machine Learning Research, 12, 2825–2830. https://jmlr.csail.mit.edu/papers/v12/pedregosa11a.html

The pandas development team. pandas-dev/pandas: Pandas [Computer software]. 10.5281/zenodo.3509134

Harris, C. R., Millman, K. J., Van Der Walt, S. J., Gommers, R., Virtanen, P., Cournapeau, D., Wieser, E., Taylor, J., Berg, S., Smith, N. J., Kern, R., Picus, M., Hoyer, S., Van Kerkwijk, M. H., Brett, M., Haldane, A., Del Río, J. F., Wiebe, M., Peterson, P., … Oliphant, T. E. (2020). Array programming with NumPy. Nature, 585(7825), 357–362. 10.1038/s41586-020-2649-2

Ansel, J., Yang, E., He, H., Gimelshein, N., Jain, A., Voznesensky, M., Bao, B., Bell, P., Berard, D., Burovski, E., Chauhan, G., Chourdia, A., Constable, W., Desmaison, A., DeVito, Z., Ellison, E., Feng, W., Gong, J., Gschwind, M., Hirsh, B., Huang, S., Kalambarkar, K., Kirsch, L., Lazos, M., Lezcano, M., Liang, Y., Liang, J., Lu, Y., Luk, C., Maher, B., Pan, Y., Puhrsch, C., Reso, M., Saroufim, M., Siraichi, M. Y., Suk, H., Suo, M., Tillet, P., Wang, E., Wang, X., Wen, W., Zhang, S., Zhao, X., Zhou, K., Zou, R., Mathews, A., Chanan, G., Wu, P., & Chintala, S. (2024). PyTorch 2: Faster Machine Learning Through Dynamic Python Bytecode Transformation and Graph Compilation [Conference paper]. 29th ACM International Conference on Architectural Support for Programming Languages and Operating Systems, Volume 2 (ASPLOS ‘24). 10.1145/3620665.3640366

Ross Wightman, Nathan Raw, Alexander Soare, Aman Arora, Chris Ha, Christoph Reich, Fredo Guan, Jakub Kaczmarzyk, mrT23, Mike, SeeFun, contrastive, Mohammed Rizin, Hyeongchan Kim, Csaba Kertész, Dushyant Mehta, Guillem Cucurull, Kushajveer Singh hankyul, … Yusuke Uchida. (2023). rwightman/pytorch-image-models: v0.8.10dev0 Release (v0.8.10dev0). Zenodo. 10.5281/zenodo.7618837

Casper da Costa-Luis, Stephen Karl Larroque, Kyle Altendorf, Hadrien Mary, richardsheridan Mikhail Korobov, Noam Yorav-Raphael, Ivan Ivanov, Marcel Bargull, Nishant Rodrigues, Guangshuo Chen, Mikhail Dektyarev, mjstevens777, Matthew D. Pagel, Martin Zugnoni, JC, CrazyPython, Charles Newey, Antony Lee, … Jack McCracken. (2024). tqdm: A fast, Extensible Progress Bar for Python and CLI (v4.66.5). Zenodo. 10.5281/zenodo.13207611

